# Virginia Accountable Health Engagement and Action Dashboard (Va-AHEAD) Protocol: Identifying Community Drivers of Health Outcomes Through Bright Spot and Community Asset Mapping

**DOI:** 10.64898/2026.11.17.26368276

**Authors:** Alex H Krist, Jacqueline B Britz, Roy T Sabo, E Marshall Brooks, Darya S Niaki, Adam J Funk, Sheila A Ward, Jennifer L Gilbert, Benjamin Webel, Marie Shoen, Jong Hyung Lee, Cynthia Burwell, Emily Zimmerman, Johnna Berryhill, Mignonne Guy, Carrie Miller, Jennifer Lambert, Meredith Young, Mark Carozza, Michael Topmiller, Derek Chapman, Scott Strayer, Patrick Nana-Sinkam, Deborah DiazGranados, Leah Gregory, Tyler Burton, F. Gerard Moeller

**Author notes:** Address correspondence: Alex Krist MD MPH. One Capital Square, Room 631 830 East Main St, Richmond VA 23219.

## Abstract

**Introduction:** Health outcomes vary substantially across communities due to multiple factors operating across the socioecological framework. Researchers, health systems, public health and government agencies, and communities often lack integrated data systems that identify communities performing better or worse than expected, characterize factors associated with outcomes, and support action. We propose to develop the Virginia Accountable Health Engagement and Action Dashboard (Va-AHEAD) to identify communities with unexpectedly favorable outcomes (Bright Spots) and the modifiable factors that contribute to their success.

**Objectives:** This protocol describes the design and methodology of a statewide, community-engaged translational science initiative to identify structural factors, community assets, and modifiable drivers associated with population health outcomes using a strengths-based approach. The project aims to co-create community health measures, identify bright spot communities and their assets, support use of findings by researchers and stakeholders, and evaluate impact.

**Methods:** Va-AHEAD integrates person-level and community-level data sources from the Virginia All-Payer Claims Database data, mortality data, census-based measures, and other socioecological datasets. Guided by a coalition of community advisory partners, the project will conduct bright spot analyses and community asset mapping to identify communities performing better than expected and the modifiable community factors that may contribute to their success across 15 priority health topics. Findings will be disseminated through an interactive dashboard, community products, and a statewide learning community. Measures of success include dashboard use, research productivity, care transformation activities, and public health and community action.

**Significance:** Va-AHEAD addresses a critical need for community-informed data systems that move beyond identifying problems to understanding factors associated with success. By integrating advanced analytics, stakeholder engagement, and dissemination strategies, this project will create a scalable model for identifying and spreading effective community approaches to improving health outcomes.

## INTRODUCTION

Health outcomes vary across individuals and communities according to socioeconomic status, geographic location, race, ethnicity, and other factors.^1^ Although differential outcomes have been recognized for more than half a century,^2,3^ significant gaps persist, and meaningful progress toward addressing them remains limited. For example, there is a ten year difference in life expectancy for Black and Native Americans compared to others.^4^ National quality measures demonstrate that compared to White people, Black people receive worse care on 76 out of 190 (40%) measures and Hispanic people on 58 out of 167 (35%) measures.^5^ Black, Hispanic, and Asian people are less likely to receive recommended preventive care and more likely to suffer worse outcomes.^6,7^ Social and structural drivers of health such as poverty, unemployment, inadequate education, and unstable housing all contribute to poor health.^8–13^ Additionally, place matters.^14,15^ The life expectancy of residents can vary by 15 to 20 years across census tracts just a few miles apart (Figure 1),^16–18^ and compared to urban communities, rural communities have higher rates of preventable conditions and unhealthy behaviors.^19^

**FIGURE 1.**
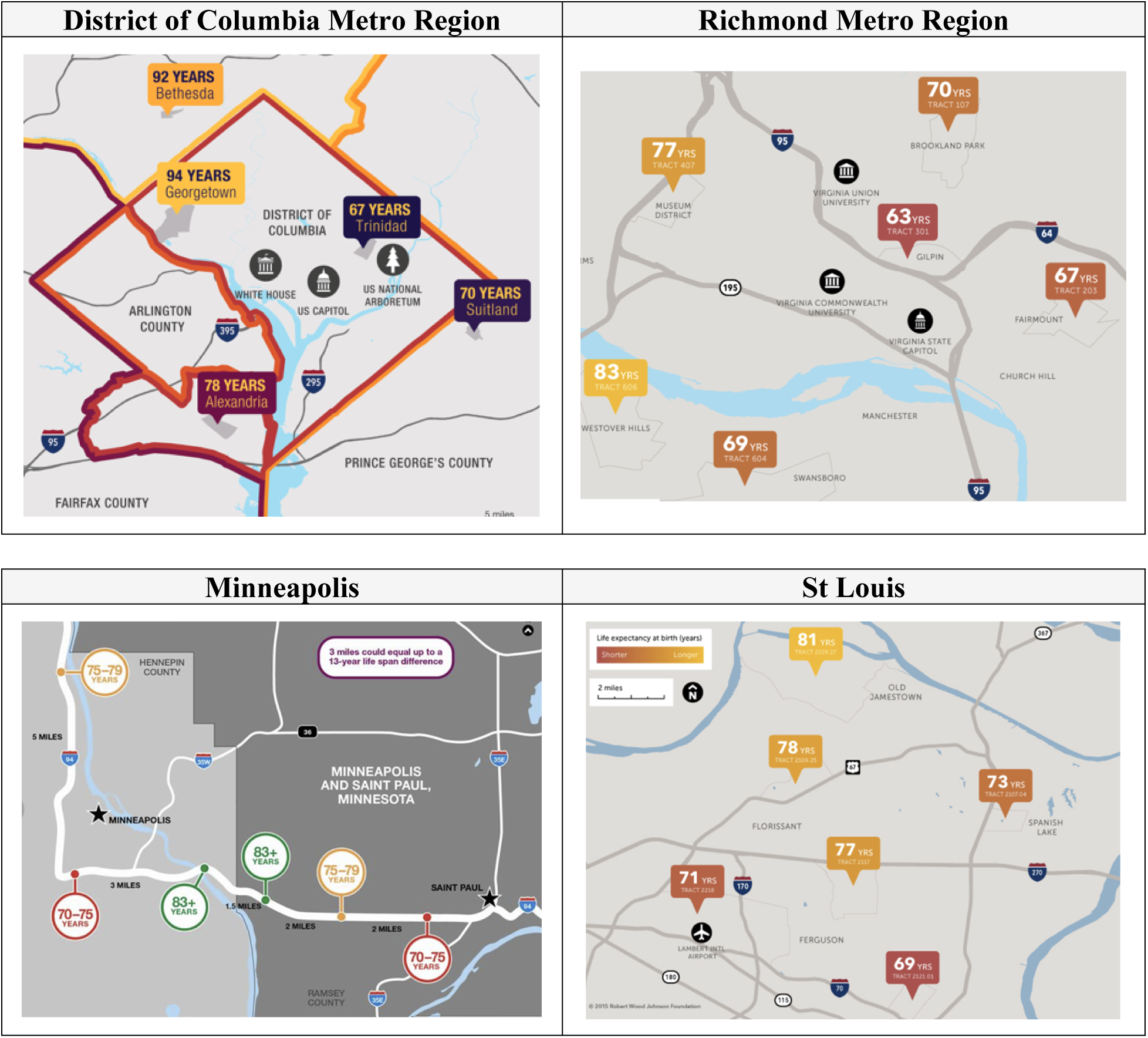
Short Distances Lead to Large Differences in Life Expectancy^44^. Maps of four U.S. metropolitan regions illustrate how life expectancy can vary substantially between communities separated by only a few miles, highlighting the importance of place in shaping health outcomes. Additional maps and project details are available at https://societyhealth.vcu.edu/work/the-projects/mapping-life-expectancy.

Health outcomes are shaped by a range of factors that affect individuals and communities in different ways.^20^ These differences are often preventable and grounded in systemic factors. Research has shown that the root causes of poor health extend well beyond healthcare, and that behaviors, social and economic factors, and the environment are the primary drivers of health and wellbeing.^21–23^ Improving health outcomes requires addressing these systemic differences across sectors through sustained, longitudinal collaboration at the local, state, and national levels.^24,25^

### The Role of Translational Science and the Roadblock

Translational science can play an essential role serving as an academic anchor, creating resources for community-driven solutions, and conducting research to inform practice and community investment to address differences in health outcomes.^26^ However, addressing multi-sector systemic differences is challenging and our understanding about how to best do this is limited.

A key barrier to this research is the lack of local data and meaningful community input needed to establish priorities, identify underlying drivers of health disparities, develop targeted interventions, and monitor progress over time. Although large population health datasets are increasingly available, they often lack community-level analyses, are difficult to access and interpret, provide limited insight into the underlying drivers of health outcomes, and rarely incorporate community perspectives. **Local data.** We know that the US has a worse life expectancy than similar developed countries and that many of the conditions driving this difference are preventable.^23,27,28^ While knowing these drivers of poor health on a macrolevel is helpful, translational science researchers need to know which communities they serve are disproportionately affected by which problems. This can prioritize what to study locally and who should be included in research. **Underlying causes.** The factors contributing to and causing different health outcomes are complex, interconnected, and span many sectors. Local data is needed across a wide range of socioecological and life-course factors. Advanced analytic methods for evaluating natural experiments (local and temporal changes from research, clinical care, community investment, or trainings), multivariate geospatial modeling, and longitudinally tracking outcomes can be harnessed to explore relationships between these many variables. This combination of data and analytics can inform the design of interventions that may have the greatest potential for success. **Community driven.** Data alone can only partially address this translational science roadblock. Deliberative multi-stakeholder engagement is also essential to setting priorities, provide local context to understand how to interpret the data, determine if associations may reflect causality, and identify potential solutions based on local values, preferences, and lived experience. Community engagement helps researchers to interpret data correctly, align data with community-identified priorities, and locally tailor potential interventions.

### Innovation

Our proposal has three elements that make it highly innovative and will transform our health outcomes research:

1. Create a community-determined system for accountability – by clearly identifying specific health outcomes and their consequences, and then linking people and communities to researchers, care teams, policymakers, and learners we will create a clear, measurable, and objective system of accountability.

2. Adopt a strengths-based approach – by focusing on bright spot communities that are thriving and assisting communities with worse health outcomes to develop similar assets and resources, we create a strengths-based and respectful roadmap for change.

3. Include the full range of socioecological variables – by considering all variables that could influence health using a combination of person-level and community-level data, we can better design interventions to address the complex topic of health outcomes.

## METHODS

### Project Overview

Funded by the National Center for Advancing Translational Sciences (NCATS; Grant RC2TR005115) through its RC2 Specialized Innovation Program (PAR-25-156), this project develops and disseminates a novel specialized innovation to support clinical and translational science, the Virginia Accountable Health Engagement and Action Dashboard (Va-AHEAD). Va-AHEAD displays community-level data generated through our novel bright spot and community asset mapping approach, which operationalizes the socioecological model by accounting for the full range of factors influencing health outcomes.^29,30^ This analytic approach integrates robust socioecological data with local context to identify community-level protective factors and potential drivers of health outcomes. The overall approach is shown in Figure 2.

**FIGURE 2.**
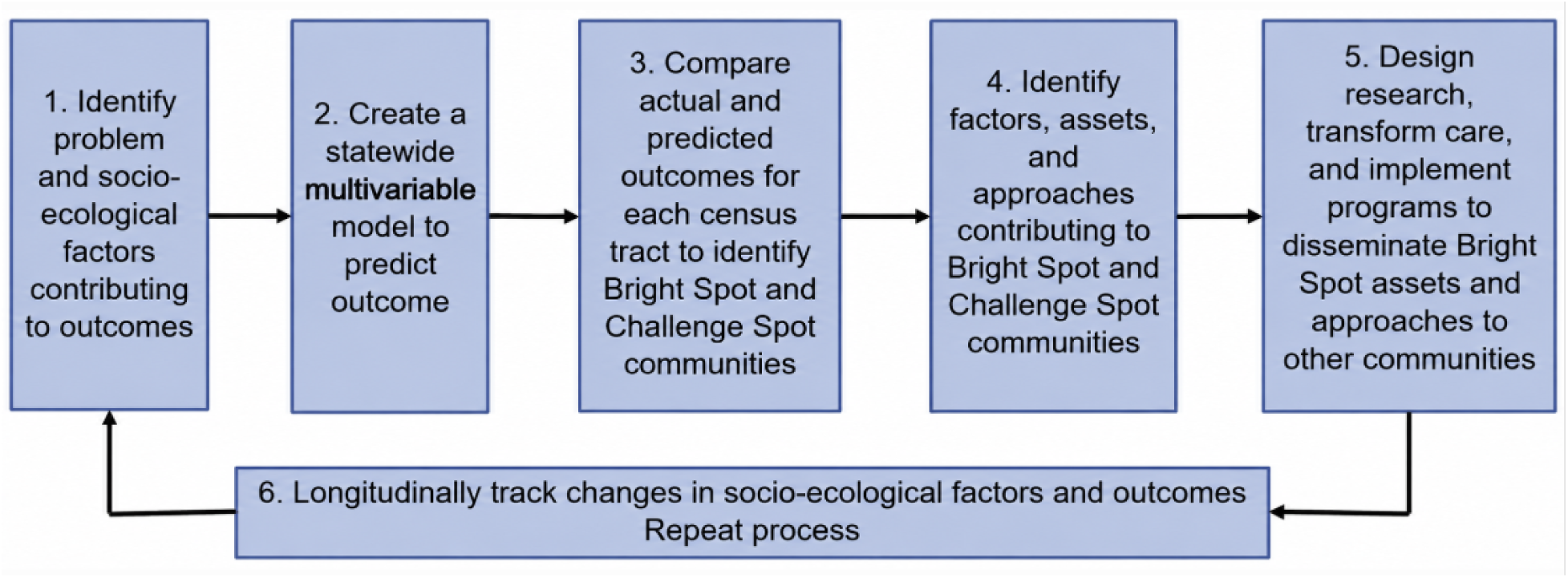
Overview of Bright spot and Community Asset Mapping Approach. Our bright spot and community asset mapping approach includes 6 steps: 1) identify the socioecological factors that contribute to outcomes, 2) conduct a multivariable analysis to create an overall model (statewide) that will predict outcomes based on socioecological factors, 3) apply overall model to each census tract and do a residual analysis to calculate the difference between actual versus predicted outcomes to identify bright spots with higher than predicted and challenge spots with lower than predicted outcomes, 4) compare characteristics of bright and challenge spot communities to identify factors, assets, and approaches that contribute to outcomes, 5) use findings to design research, transform care, and implement programs that will disseminate bright spot resources to communities in need, and 6) longitudinally track the health outcomes and contributing socioecological factors for each community.

Throughout the process, we will co-create content with a broad and representative cadre of community partners from across the state. Va-AHEAD will address 15 community identified health priorities (2-4 added per year) and will include community level measures, analytic findings, and tools and products developed by the community. Content will be annually updated and displayed longitudinally. The project consists of seven phases: (1) prioritize and co-create health outcome goals and measures with communities, (2) identify initial bright spot communities that perform better than expected and factors that may contribute to their success, (3) conduct community asset mapping to understand the factors that create bright spot communities, (4) refine the bright spot model to further inform interventions, (5) develop shared meaning, products, and action from the data, (6) disseminate and support use of content, and (7) measure the impact of the approach, change in health outcomes, and evolution of partnerships.

### Project Aims

We define success of this project as an increase in impactful health outcomes research, clinical and community care transformation, and state programs; inclusion of more broad and representative people and communities in research; and dissemination of assets and approaches used by bright spot communities to other communities. Project specific aims include:

### Aim 1 (Creation)

Engage a broad range of community partners to co-create the Va-AHEAD measures and analyses. Meet with community partners weekly to monthly to address 15 health outcomes topics. For each topic, we will identify factors that contribute to outcomes, conduct a bright spot and community asset analysis, and design dashboard measures and products. Annually, we will assess the impact of the process on our relationship with community partners.

### Aim 2 (Use)

Disseminate and support researchers, care teams, state and national programs, community partners, and learners to use Va-AHEAD. Partners will include over seven community advisory boards, dozens of community partner organizations, three universities (VCU, Eastern Virginia Medical School (EVMS), and Norfolk State University (NSU)), four health systems (VCU, Inova, Sentara, and Ballad), and state and national programs (Virginia and city/county departments of health, Department of Medical Assistance Services). From the Va-AHEAD database, support requests, field records, training records, and Annual Partnership Survey, we will assess the number of people and organizations supported to use the new content and how content is used.

### Aim 3 (Effectiveness)

Demonstrate that use of Va-AHEAD and its products and findings increase research and training, change care, and affect state and national programs to improve health outcomes, and that these interventions extend bright spot assets to communities in need. We will use field notes, Annual Partnership Survey, Group Level Assessments, annual and final research reports, and Va-AHEAD data to determine the (a) frequency and perceived impact of dashboard use, (b) number and impact of studies, care transformations, policies, and teachings, and (c) extension of bright spot approaches and assets to communities with the greatest health needs.

### Ethics statement

The study will be conducted in accordance with US Code of Federal Regulations (45 CFR Part 46) and ICH Good Clinical Practice (ICH GCP) E6(R2). The protocol was reviewed by the Virginia Commonwealth University (VCU) Institutional Review Board (IRB) and deemed not human subjects research (IRB File #HM20031637).

Data sources to create community health metrics or existing community measures used for our bright spot analyses are all de-identified. Meetings with community members will be recorded. Discussions will focus on the priorities and perspectives of the communities they represent and will not include discussions about personal health information or other protected information. Participants will be informed about the purpose of study and nature of discussions prior to participation and will have the opportunity to withdraw from discussions and participation as desired.

### Setting

The study will include every census tract, county, and city in Virginia, with the geographic unit varying according to the available data and topic assessed. Virginia is a geographically and demographically representative state that includes urban, suburban, rural, coastal, Appalachian, and medically underserved communities. Virginia’s heterogeneity in population characteristics, health systems, socioeconomic conditions, and community contexts closely reflects the heterogeneity seen across many U.S. states, making it an ideal setting for developing and evaluating community health outcomes.

### Phase 1: Community Prioritization and Co-Creation

Community engagement is foundational to Va-AHEAD and occurs through three complementary structures: the Community Data Translation Team (CDTT), Topic Teams, and dissemination opportunities. Community members serve as equal partners throughout the research process, helping to establish priorities, interpret findings, develop dissemination products, and guide implementation. We assembled the CDTT at the outset of the study to provide overall governance and ensure that Va-AHEAD reflects priorities from across the Commonwealth. Members were selected to broadly represent Virginia’s health interests and were drawn from established community partnerships of the Wright Center for Clinical and Translational Research and VCU. The CDTT meets monthly throughout the five-year project and is responsible for selecting the health topics addressed through bright spot and community asset mapping, identifying Topic Team members, serving as consultants throughout each assessment, supporting dissemination activities, and providing ongoing oversight of the project’s scientific and community-engaged direction.

For each priority area, we assembled 7 Topic Teams to oversee the 15 bright spot and community asset mapping assessments. The topics identified by the CDTT include primary care, substance use, mental health, cancer, adverse childhood events, obesity, and cultural arts. Topic Team members were identified with guidance from the CDTT using a snowball sampling approach to recruit key stakeholders from across Virginia with relevant expertise and lived, professional, and organizational experience. Topic Teams meet 6–12 times over the course of each assessment and are responsible for identifying meaningful outcomes, selecting candidate factors for bright spot analyses, interpreting findings, guiding community asset mapping, refining analytic models, developing dashboard and dissemination content, and supporting dissemination. Collectively, the CDTT and Topic Teams include more than 75 members representing over 63 organizations distributed throughout Virginia, including clinicians, patients, public health, community organizations, government, philanthropy, advocacy organizations, and community members, ensuring that the project reflects broad perspectives from across the Commonwealth. Biographies of CDTT and Topic Team members are available at https://virginia-ahead.org/.

### Phase 2: Identify Bright Spot Communities

The identification of bright spot communities is shown in steps 1-3 of Figure 2. In our approach, each community is characterized along two dimensions: (1) observed outcomes and (2) performance relative to predicted outcomes. **Hot spots and cold spots** represent communities with higher and lower observed outcomes, respectively. **Bright and challenge spots** represent communities performing greater and lower than predicted based on their structural factors and key drivers.

For each topic, the Topic Team identifies one to three outcomes that are meaningful to communities and identifies structural factors and key drivers that may influence these outcomes. Each outcome serves as the dependent variable for a bright spot analysis, with structural factors and key drivers serving as independent variables. Variable selection is informed by evidence reviews, Topic Team members’ knowledge of the communities they represent, and the socioecological and life-course frameworks (Table 1).^23^ The socioecological framework accounts not only for the proximal influences of healthcare and behavior on biological outcomes, but also the upstream influences of socioeconomic status and social and physical environments.^11,12^ Life-course research recognizes health as a longitudinal process, influenced by interacting social, environmental, and biological factors that shape health trajectories over the lifespan and temporal health trends in communities.^31^ The Data and Analysis Team supports variable selection by identifying appropriate census tract-level measures from the Virginia All-Payer Claims Database (APCD), Virginia Department of Health mortality data, HealthLandscape Vital Signs library, and additional data sources as needed.^32,33^ Variables must represent directly observed or measured characteristics rather than synthetic estimates derived from population demographics. For example, small-area estimates of health behaviors that are modeled using age, sex, race/ethnicity, or other population characteristics are not used as they would merely reflect the demographics of a community

**TABLE 1.** Examples of Structural Factors and Key Drivers.

| **Structural Factors** | **Key Drivers** |
|---|---|
| • **Demographics.** Proportion of residents: Black, Hispanic, <5 years, >65 years, foreign born<br>• **Rurality.** RUCA classification or population density<br>• **Socioeconomic conditions.** Proportion of residents: under 200% poverty level, with high school degree or less, not working, with disability, receiving SNAP benefits or receiving public assistance<br>• **Household conditions.** Proportion of resident divorced or proportion single-parent households<br>• **Insurance coverage.** Proportion residents uninsured, with Medicaid<br>• **Transportation.** Proportion of residents without access to vehicle<br>• **Housing.** Proportion households spending >50% income on housing, eviction filing rate, public housing units<br>• **Food environment.** Proportion of residents within 1 mile of grocery store<br>• **Residential segregation.** Racial and economic segregation calculated from Index of Concentration at the Extremes<br>• **Community disadvantage.** Adjusted area deprivation index or social deprivation index or social vulnerability index or index of deep disadvantage<br>• **Community advantage.** Child opportunity index | • **Prevalence of health conditions.** Mental health conditions, substance use disorders, diabetes, and other chronic conditions<br>• **Access to care.** Spatial access to primary care, behavioral health, specialty care, and treatment facilities<br>• **Use of care.** Primary care visits, mental health visits<br>• **Screening and early detection.** Colorectal cancer screening, behavioral health screening, and stage at cancer diagnosis<br>• **Treatment and prescribing.** Opioid prescribing, medications for opioid use disorder, and guideline-recommended treatment<br>• **Quality of care.** Comprehensiveness of primary care, continuity of care, preventive care delivery<br>• **Health behaviors and community resources.** Tobacco use, physical activity, healthy food access, and availability of community-based services |

Once outcomes and independent variables are identified, the Data and Analysis Team conducts a statewide multivariable regression analysis at the census tract level to predict outcomes based on structural factors and key drivers. Model selection is guided by the distribution and measurement characteristics of each outcome, with generalized linear models used as appropriate for continuous, count, or rate-based outcomes, and Poisson regression for count data. Depending on data availability and the research question, models may incorporate multiple years of observations, interactions with time or other community characteristics, and methods to account for spatial autocorrelation. The statewide model results are applied to each census tract to calculate residuals as the difference between observed and predicted outcomes. Bright spots are defined as the 30–50 census tracts with the most favorable outcomes relative to predictions, and challenge spots as the 30–50 with the least favorable outcomes relative to predictions. Because structural factors and key drivers identified by Topic Teams may be conceptually related, the Data and Analysis Team also assesses multicollinearity among independent variables. When substantial collinearity is identified, the team conducts sensitivity analyses using alternative models in which collinear variables are entered separately. Decisions about which measures to retain are made jointly with Topic Teams, considering substantive importance, stability of associations, and model performance.

### Phase 3: Conduct Community Asset Mapping

From the initial hot spot and bright spot analyses the community asset mapping process identifies the modifiable assets, approaches, and resources that may explain why communities experience different outcomes. The process combines quantitative data with qualitative approaches to understand local context, knowledge, and perspectives. Ultimately, strengths identified through this process can be disseminated and adapted for use by other communities.

First, results from the initial bright spot multivariable analysis are shared with the Topic Team to identify **where** outcomes differ across Virginia. We create eight maps and two data tables. Four domains are displayed at two geographic levels, city/county and census tract: (1) distribution of hot and cold spots across all communities, (2) 30 to 50 highest- and lowest-ranking hot and cold spots, (3) distribution of bright and challenge spots across all communities, and (4) 30 to 50 highest- and lowest-ranking bright and challenge spots. The accompanying tables report the outcome, structural factor, and key driver values for each census tract and city/county. City/county analyses correspond to the geographic level at which many policy, programmatic, and investment decisions are made, whereas census tract analyses reveal variation within localities and identify neighborhoods where action may be needed or strengths-based lessons can be learned. In reviewing the data, Topic Teams first assess the external validity of the findings by determining whether the results are consistent with their knowledge and understanding of the communities represented.

Second, we examine **what** potentially modifiable factors may contribute to these differences. Topic Teams identify additional socioecological factors beyond the structural factors and key drivers included in the initial bright spot multivariable model, emphasizing factors that may be amenable to intervention (Table 1). When data are available for these factors, the Data and Analysis Team summarizes these measures for each census tract and city/county, and Topic Teams qualitatively compare patterns across communities to identify factors that may further explain differences in outcomes. Topic Teams then identify key stakeholders from a range of bright, challenge, hot, and cold communities for interviews by the Community Engagement Team. Interviews explore stakeholders’ knowledge and perspectives regarding local factors, assets, resources, and approaches that may contribute to community outcomes, including their perceptions of how these factors influence outcomes and insights into potentially important factors for which quantitative data are unavailable.

Third, the Data and Topic Teams examine **for whom and under what circumstances** these modifiable factors may differ. We identify community characteristics that may modify these relationships, such as rurality or level of social deprivation, and conduct quantitative subgroup analyses and stakeholder interviews to determine whether communities sharing these characteristics have similar or different assets, challenges, and modifiable factors. For example, factors associated with better-than-predicted outcomes in rural communities may differ from those in urban communities, or approaches that appear important in communities with high social deprivation may differ from those in more advantaged communities.

Finally, we integrate findings across the bright spot analysis, additional quantitative comparisons, subgroup analyses, and stakeholder interviews to identify the estimated associations and community-perceived contributions of assets, resources, and approaches to outcomes. These findings are used to develop **tailored sets of potential research, clinical care, public health, community, policy, and programmatic approaches** that reflect communities’ structural characteristics, modifiable factors, assets, and needs.

### Phase 4: Refine Model

Findings from community asset mapping are used to refine the initial bright spot model. The Data and Analysis Team and Topic Team review additional factors identified through quantitative comparisons, subgroup analyses, and stakeholder interviews and determine whether any represent structural factors or key drivers that should have been included in the initial statewide model. Factors are considered for inclusion when they are conceptually related to the outcome, supported by the quantitative or qualitative findings, considered important by the Topic Team, and meet the data requirements described in Phase 2. Selected factors are added to a refined multivariable model, which is reapplied to all census tracts to recalculate predicted outcomes and residuals and determine whether the identification or ranking of bright and challenge spots changes.

Importantly, the purpose of model refinement is not to statistically adjust away the potentially modifiable assets, resources, and approaches that community asset mapping is intended to discover. Variables representing specific community programs, resources, practices, or other potentially transferable approaches are generally treated as explanatory findings rather than adjustment variables in the primary bright spot model. When a newly identified factor could reasonably function as either a key driver of the outcome or a potentially transferable community asset, we compare alternative model specifications with and without the factor. Changes in model coefficients, predicted outcomes, residuals, and bright spot classification are examined and interpreted with the Topic Team. This approach preserves the model’s ability to account for important underlying community characteristics while retaining the assets and approaches that may explain how communities achieve better-than-predicted outcomes.

### Phase 5: Develop Shared Meaning, Products, and Action (VA-AHEAD Dashboard Design)

The overarching goal of the bright spot model and community asset mapping approach is to identify community assets, resources, and approaches associated with favorable health outcomes and make these findings actionable for researchers, clinical teams, community partners, learners, government and public health officials. To do this, the Topic Teams will assemble key information for each of the seven topic domains into the Va-AHEAD website, https://virginia-ahead.org/. Content will include four major components: (1) data and measures, (2) tools and resources, (3) supported projects and partners, (4) success stories, and (5) team members. Throughout, we will use human-centered design principles to ensure content is accessible, understandable, and usable.^34^

## Data and Measures

For each topic domains, the Va-AHEAD data dashboard will display findings for each bright spot model developed by the Topic Teams in three ways: (1) interactive maps identifying hot, cold, bright, and challenge spots at both the city/county and census tract levels; (2) graphs displaying each community along hot versus cold and bright versus challenge characteristics; and (3) community scorecards reporting the outcome measures, structural factors, key drivers, and additional quantitative measures identified through community asset mapping. Users will be able to filter and compare content by geographic region and community characteristics including rurality, poverty, social deprivation, and racial/ethnic composition. City/county-level findings can help guide public health programs, policies, and regional investments, while census tract-level findings can identify neighborhoods where resources and interventions may be needed or where community assets and approaches could inform action elsewhere. Measures will be updated annually to track changes over time and all data will be downloadable.

### Tools and Resources

Va-AHEAD will provide tools and resources to help users understand Bright spot findings, identify evidence-based approaches to improve outcomes, and apply lessons learned from successful communities. Features will include data export, geographic and temporal comparisons, additional analytic tools, training materials, and topic-specific reports documenting the seven phases of the assessment process. Each topic will also include a curated collection of evidence-based interventions, community resources, and promising approaches relevant to the factors and opportunities identified through bright spot and community asset mapping. These collections will summarize supporting evidence, implementation considerations, and links to relevant programs, organizations, and tools. To reach a broad range of audiences, we will develop infographics, one-page summaries, issue briefs, interactive StoryMaps, and other communication products.

### Success Stories

Va-AHEAD will feature stories of bright spot communities highlighting the resources, assets, and approaches that may explain their better-than-expected health outcomes. Stories will describe the community context, successful programs and approaches, partners involved, and lessons learned, with links to supporting evidence, implementation resources, and individuals or organizations with relevant expertise. These examples will help other communities identify promising strategies, connect with experienced partners, and consider opportunities for local adaptation.

### Supported Projects and Partners

Va-AHEAD will highlight projects and partnerships that use bright spot and community asset mapping findings to guide research, community planning, public health, and interventions to improve health outcomes. Each project will include a summary of its goals, partners, activities, findings, and relevant contacts or resources. These examples will demonstrate how Va-AHEAD findings are applied and facilitate collaboration and replication of promising approaches.

### Phase 6: Disseminate and Support Use of Content

Our approach to disseminating Va-AHEAD findings and supporting their use will be guided by the Agency for Healthcare Research and Quality’s *Dissemination Planning Tool* framework, which emphasizes identifying dissemination content, target audiences, partners, and communication strategies, as well as evaluating and adapting dissemination efforts over time.^35^ This systematic approach will help ensure that findings reach intended audiences and are used to inform research, policy, practice, and community action.

### Partner-Based Dissemination

Va-AHEAD has an established network of dissemination partners, including participating universities (VCU, EVMS, NSU), health systems (VCU Health System, Inova, Sentara, Ballad), translational science partners (VCU Wright Center for Clinical and Translational Research), seven established community advisory boards, and dozens of community organizations. Topic Teams will also identify and engage topic-specific dissemination partners, such as the Virginia Ambulatory Care Outcomes Research Network (ACORN) for primary care, the Virginia Opioid Abatement Authority for opioid-related findings, and the Virginia Colorectal Cancer Care Alliance for colorectal cancer. We will work with these partners to share findings through multiple communication channels, including direct outreach, partner communications, webinars, conference presentations, media, policy briefs, and social media. Training videos and other educational materials will help users understand and apply dashboard findings.

### Support and Use Consultations

Users will be able to request consultations for more intensive assistance applying Va-AHEAD data and findings. Support may include developing new measures or analyses, adapting or creating products for specific audiences, accessing underlying data, obtaining biostatistical or community engagement expertise, generating pilot data for grant applications, and developing new projects or funding proposals. Through the VCU Wright Center for Clinical and Translational Research, our team will provide up to six hours of consultation at no charge. More extensive assistance may be supported through grants, foundation funding, or institutional partnerships.

### Phase 7: Measure the Impact

We will assess three dimensions of impact corresponding to our study aims: (1) creation of Va-AHEAD content through community partnerships, (2) dissemination and use of Va-AHEAD, and (3) effectiveness in supporting interventions that extend bright spot assets and approaches to communities with identified needs. Table 2 summarizes the outcomes, data sources, and analytic approaches for each aim.

**TABLE 2.** Overview of RC2 Program Outcomes and Evaluation Plan.

| Aim | Outcomes | Data Sources | Analytic Plan |
|---|---|---|---|
| **Aim 1 (Creation)** co-create Va-AHEAD with community partners | • Number of Va-AHEAD measures and products<br>• Bright spot findings (socioecological factors identified, hot/cold and bright/challenge spot communities, factors contributing to outcomes, potential solutions)<br>• Process of community co-development (who is engaged, how they are engaged, how partnership changes content)<br>• Impact of partnership on community relationships | • Va-AHEAD database<br>• Meeting and field notes<br>• Annual partnership assessment | • Baseline bright spot and community asset mapping analysis for the 15 topics<br>• Partnership process with community (qualitative analysis)<br>• Annual change in collective capacity building score (generalized linear mixed models) |
| **Aim 2 (Use)** disseminate and support use of Va-AHEAD | • Number of Va-AHEAD users and communities they represent<br>• Va-AHEAD content used and frequency of use<br>• Number and types of research, care, policy, and teaching interventions supported | • Dissemination and training attendance<br>• Va-AHEAD database<br>• Va-AHEAD user survey<br>• Annual partnership assessment | • Number of Va-AHEAD users, Va-AHEAD content used, and interventions and communities supported (descriptive statistics)<br>• Change in academic output from efforts (grants, funding, interventions, presentations, papers, and trainings) |
| **Aim 3 (Effectiveness)** demonstrate Va-AHEAD use increases interventions and improves outcomes | • More interventions that address the 15 prioritized topics and use identified solutions<br>• More challenge spot communities participate in research, receive care, benefit from community programs, or are trained<br>• Extension of bright spot assets and resources to communities with identified needs<br>• Improved outcomes | • Va-AHEAD database<br>• Annual and final project research reports<br>• Annual partnership assessment | • Reach and characteristics of communities and people included (descriptive statistics)<br>• Increase in resources for communities (generalized mixed models)<br>• Improvement in socioecological factors and outcomes for challenge spot communities (generalized linear mixed models) |

### Data Collection

We will use five complementary data sources to assess project outcomes: (1) Va-AHEAD database, including community measures, bright spot analyses, and website utilization; (2) meeting minutes, recordings, transcripts, and field notes documenting community engagement and supported projects; (3) dissemination and training attendance records; (4) annual and final reports from Va-AHEAD-supported projects; and (5) annual partnership assessments. The partnership assessments, conducted with participating health systems, the CDTT, Topic Teams, and community organizations, will include surveys and follow-up interviews with representatives of each organization. Surveys will use the E2E Framework to assess partnership functioning, trust, community capacity, and readiness for collective action.^36–39^ Interviews will explore the nature and impact of partnerships, shared projects, communities reached, and resulting products and outcomes. Together, these sources will allow us to assess how Va-AHEAD is developed, used, and translated into community action.

### Outcomes

For Aim 1 (Creation), we will assess the number and types of measures, bright spot models, community asset mapping findings, tools, and dissemination products developed. We will also evaluate the community co-development process, including participation, contributions to dashboard content, and changes in partnership capacity and community mobilization scores over time. For Aim 2 (Use), we will measure the number and characteristics of researchers, clinicians, policymakers, learners, organizations, and communities reached; frequency and types of dashboard use; and the number and types of research, care delivery, public health, policy, and training activities supported. We will also assess academic and programmatic outputs, including grants, funding, interventions, presentations, publications, and workforce training, comparing productivity during the project with the five years preceding the project.

For Aim 3 (Effectiveness), we will assess whether Va-AHEAD increases interventions that address community-identified health priorities, expands participation of underserved communities in research and programs, and supports the adoption of resources, assets, and approaches identified in bright spot communities. **Our primary outcome is the increase in bright spot assets and resources implemented in challenge spot communities.** We will use project-supported reports and annually updated community measures to determine whether these approaches are being extended to communities with identified needs. Our secondary outcome is improvement in community-level structural factors, key drivers, and health outcomes. Because meaningful changes in population health may require more than five years, we will assess these outcomes longitudinally while recognizing that some impacts may extend beyond the study period.

### Quantitative Analytic Plan

We will use descriptive statistics to summarize dashboard content, users, dissemination activities, supported projects, and academic outputs. Annual bright spot analyses will assess changes in community characteristics, observed and predicted outcomes, and bright spot classifications over time. We will use generalized linear mixed models to evaluate increases in supported interventions, alignment of projects with identified community needs, inclusion of underserved populations, extension of bright spot assets and resources to challenge spot communities (primary outcome), and improvements in community health outcomes (secondary outcome). Analyses will be conducted at the census tract level, with fixed effects for time, relevant area-level characteristics from the American Community Survey, and area-level random effects to account for spatial correlation and repeated measurements. Outcomes will be modeled as continuous or count measures, as appropriate, and nonlinear trends will be explored using polynomial or spline terms.

### Qualitative Analytic Plan

We will analyze meeting transcripts, field notes, and annual semi-structured interviews to understand the processes of co-developing, disseminating, and applying Va-AHEAD findings. We will use a combination of template-based coding informed by the Consolidated Framework for Implementation Research and emergent coding to identify themes specific to the project.^40–42^ Analyses will examine how community partnerships influence dashboard content, interpretation of findings, development of interventions, dissemination, and community action. We will also identify disconfirming evidence and integrate qualitative findings with quantitative outcomes to better understand the mechanisms and contextual factors contributing to project impact.

### Power Estimation

For each bright spot assessment, our primary outcome is the number of identified bright spot assets and resources implemented in challenge spot communities. Among the 30 communities with the most severe worse-than-predicted outcomes, assuming no identified assets or resources at baseline and a standard deviation of two, we estimate greater than 80% power to detect an average increase of one asset or resource over time. Pooling approximately 500 challenge spot communities across the 15 planned assessments provides at least 80% power to detect an average increase of 0.25 assets or resources. Improvements in community health outcomes will be assessed as secondary outcomes, recognizing that these changes may require longer follow-up.

## DISCUSSION

Va-AHEAD creates a lasting translational science resource to guide and inform research, clinical and community care; public health programs; local, state, federal government action; and training. It also presents a model that can be replicated for other health topics and in other geographic areas. By integrating comprehensive socioecological data with bright spot analyses and community asset mapping, Va-AHEAD moves beyond identifying differences in health outcomes to understanding the resources, assets, and approaches that help communities achieve better-than-expected outcomes. The resulting community scorecards provide a sustainable, data-driven roadmap for improving health outcomes, identifying where needs are greatest, what factors contribute to those needs, and the potentially modifiable approaches that may improve outcomes. Grounded in the full socioecological framework and informed by community partners, these scorecards help researchers, care teams, government agencies, and community organizations prioritize investments, design interventions, and track progress over time.

Once created, Va-AHEAD and its bright spot analyses are sustainable beyond the grants project period. License fees for the Va-AHEAD ArcGIS platform and datasets are supported through the Wright Center for Clinical and Translational Research. We use standardized formats to store, access, and analyze data sources. As we develop new measures and bright spot analyses, we maintain code, scripts, and instructions in a GitHub repository (https://github.com/VCUWrightCenter/Va-AHEAD) to streamline annual updates and facilitate replication and use by others. We anticipate that continued engagement of researchers, local and state government, care teams, community organizations, and learners will generate new opportunities to sustain and expand Va-AHEAD. Together, these resources establish an enduring infrastructure for monitoring community health, identifying opportunities for improvement, and evaluating the impact of interventions and investments.

Our approach has three main limitations. First, although Virginia’s APCD includes approximately 5.3 million of the state’s 8.2 million residents, it excludes some veterans, uninsured individuals, and commercial claims.^43^ However, the APCD includes complete Medicare and Medicaid claims, populations particularly important for understanding differences in health outcomes. Race and ethnicity are available for approximately 60% of claims and are supplemented by community characteristics from census data. We address incomplete claims through adjustments for missing commercial data and by emphasizing comparisons across geographic areas and trends over time. Second, mortality data may contain inaccuracies, particularly in the assignment of causes of death, although these errors are not expected to systematically bias geographic or temporal comparisons. Third, our long-term goal of improving health outcomes, particularly for people and communities with worse outcomes, is ambitious and requires sustained effort and cross-sector collaboration. Meaningful changes may extend beyond the five-year study period. Nevertheless, Va-AHEAD builds on established infrastructure and partnerships and provides the data, tools, and community engagement processes needed to establish priorities, guide action, and measure progress.

If successful, Va-AHEAD will support an expanding range of research, clinical and community care, policy, and training initiatives. We anticipate that these efforts will become increasingly sophisticated and impactful as we learn from our findings, expand the range and geographic distribution of partners and projects, and continue to update and adapt Va-AHEAD. We also plan to extend Va-AHEAD maps to other states as a proof of concept for expansion beyond Virginia. The resources and findings developed through this project provide a foundation for replication across other Clinical and Translational Science Award Program Hubs. The underlying data sources, including state mortality records, all-payer claims databases, and the American Community Survey, are available in many states, and our standardized data architecture, analytic code, and instructions facilitate adaptation elsewhere. During project years 3–5, we will engage other CTSA hubs through established partnerships, national meetings, and publications, and work with NCATS leadership to explore mechanisms for supporting broader dissemination and replication.

## CONCLUSION

Ultimately, Va-AHEAD seeks to establish a sustainable, community-driven learning system that connects data to action. By identifying what makes communities successful, translating those findings into practical resources and interventions, and tracking changes in community health over time, Va-AHEAD provides a replicable approach for directing research, care delivery, and public health investments, toward improving health outcomes for all communities.

## Data Availability

No datasets were generated or analyzed for the current study protocol. All data generated through the Virginia Accountable Health Engagement and Action Dashboard (Va-AHEAD) project will be made publicly available and downloadable without restriction at https://virginia-ahead.org/. Data definitions and executable scripts used to generate study variables and conduct analyses will be publicly available at https://github.com/VCUWrightCenter/Va-AHEAD. Data and code will be released as they become available throughout the project.

